# Experienced inclusion and recognition amongst people with spinal cord injury: A comparative study in Norway, The Netherlands, and Australia

**DOI:** 10.1101/2024.06.16.24308999

**Authors:** Annelie Schedin Leiulfsrud, Kristian Bernhof Ellinggard, Marcel W.M. Post, Mohit Arora, Håkon Leiulfsrud

## Abstract

**Background:** The aim of this article is to study inclusion and recognition experienced amongst people with a Spinal Cord Injury (SCI) in Norway, The Netherlands, and Australia. This is approached both from the perspectives of an interest in the impact of mobility limitations versus social attitudes, and from a consideration of differences between societies.

**Methods:** The data derive from the core questionnaire of International Spinal Cord Injury Community Survey with extended national modules on the attitudes and values of respondents from Norway, The Netherlands, and Australia. The data gathered in 2017-18 include 2,450 participants aged 18 years or older. The data are analysed and presented with descriptive statistics and OLS regression analyses. In order to explore our main questions, we run regression controlling for country effects in addition exploring within country effects.

**Results:** Mobility limitations are a substantially weaker predictor of self-perceived inclusion and recognition than experiences of negative attitudes towards disabled people. Stereotypical attitudes and norms in society are shown to have various impacts on inclusion in the three countries. The Norwegian respondents report overall better results on inclusion and recognition than respondents in Australia and The Netherlands, illustrating the importance of national contexts.

**Conclusions:** Challenges associated with inclusion, recognition, and respect after SCI need to be defined in a language broader than mobility limitations and stereotypical attitudes towards people with an SCI. The main road to both inclusion and recognition in society is primarily linked to job and educational status, in addition to family and friends. The results are of particular interest as measures to support reintegration into society, including a strengthening of labour market integration programs.

## Introduction

This article aims to study inclusion and recognition experienced amongst people with a Spinal Cord Injury (SCI) in Norway, The Netherlands, and Australia. In particular, we probe the relative impact of 1) mobility limitations and 2) negative societal attitudes about disabled people and how study participants experience inclusion and recognition. In addition, the cross-national research design enables us to study national, as well as more general features of social inclusion and social recognition.

In this article we introduce two types of constraints with a potential impact on self-perceived inclusion and recognition. The *first* type of constraint is mobility limitations. Mobility limitations may hamper moving around and access to public and private buildings but is also linked to individual identity management and performance [1–3]. Since use of a wheelchair or other highly visible mobility aids in public may represent a social stigma or sign of disability and exclusion [2,4] it may be wise to distinguish between wheelchair users and those able to walk with or without visible markers, such as crutches or canes. This is also an assumption in congruence with previous research from The Netherlands, showing significant associations between categories of greater mobility level (using an electric wheelchair, using a manual wheelchair, walking with aids or walking without aids) with higher levels of overall level of well-being [5]. The *second* type of constraint found in the disability literature stresses more general negative societal attitudes towards disabled people in society as a strong predictor of social exclusion [6]. The two constraints lead us to an assumption that mobility constraints directly (constraint 1) or indirectly through negative attitudes in society (constraint 2) lead to a diminished sense of social inclusion. To learn more about how these constraints are linked to perceptions of being an active and valuable member of society may help us to gain a better understanding of how the social mechanisms at play impact individual and collective perceptions of inclusion and recognition in different societies.

### Mobility constraints as a marker and potential obstacle to social trust and social inclusion

Mobility limitations as an inclusion factor in traditional rehabilitation literature tend to be described in negative terms of a need of help, powerlessness, life space limitations and associated with bodily impairment [7]. This is also shown in the description of those who are wheelchair-bound but supplemented with a distinction between active wheelchair users as more empowered and socially included, and passive wheelchair users as more depowered and socially excluded [7]. British disability researchers, Sapey, Stewart and Donaldsen [8], departing from a social model analysis of disablement, have described the wheelchair as both a liberator and an obstacle preventing full participation. The participation limitations in the British study [8] are both illustrated by the nature of the physical environment, and of perceived negative attitudes. Both of these studies are good illustrations of different narratives and problem foci associated with being wheelchair-bound, but none of them problematises potential differences between different types of mobility limitations – including distinctions between those able to move around more or less independently and those fully dependent on others. Nor does it differentiate between those who sit in a manual wheelchair and those in an electric wheelchair.

The first distinction is of potential interest as it is linked to a narrative concerning those able to move around more or less independently and those fully dependent on others. The second distinction is of interest as it may differentiate between those who can manage with their own will and body powers to get around, and those who are more dependent in terms of assistance. These aspects of mobility limitations are of interest as they relate to how people with a physical disability perceive themselves, believe others are experiencing them, or answer to questions of social trust in others, and society more broadly.

### The relevance of societal factors in explaining social trust and social inclusion

In contrast to much of previous research on social trust, social policy, and welfare or disability policy with an emphasis on more systemic features of society, our approach is to look more closely into social inclusion as “lived experience”, i.e., adopting a focus on experiences of inclusion and recognition in everyday life interactions with other people and in society. Our focus on social recognition struggles and empowerment is in line with a growing body of research with a focus on social recognition as the basis of social order and inclusion in society [9–10]. From this perspective, being recognised by others opens the door to participating fully and being included in society.

Conversely, lack of recognition by others may be seen as a form of asymmetrical power relations and an essential component of the process of social marginalisation and social exclusion [9, 11]. Social recognition of others is not *a priori* a matter of positive or negative attitudes, and as illustrated by a study by Bollier et al. [12] in Australia in which a substantial part of respondents end up answering, “I dońt know” on attitudes towards people with a disability. This is both a reply that may be interpreted as genuine uncertainty or fear of being ideologically biased; it is also a way of answering, revealing that social recognition potentially can be partial, ambiguous, or contradictory.

The theories of Honneth and Fraser [9, 11] depart from an interest in individuals as well as the institutions and social fabric of society. This is also of importance in a comparative study of how perceptions of oneself and others as well as the social climate for disabled people are played out in different societies. For our purpose, bringing societal factors into the analysis of experienced mobility contraints may also been seen as an important mediator.

### Inclusion and social trust in a comparative perspective

On paper, Norway, The Netherlands, and Australia share several similarities that may play out in an analysis of perceptions of inclusion amongst people with SCI. All three societies are based on overall high levels of social trust in fellow citizens, institutions, and society; social trust is generally used as a proxy for high levels of social cohesion and social inclusion in society [13] All three societies have well-developed social security systems in terms of health, pensions, and unemployment [14–16]. The employment and relative poverty rates for people with a disability and the threshold for receiving disability-related benefits are, however, substantially worse in Australia than in either The Netherlands or Norway [17]. This may suggest a harsher social and economic climate in Australia for disabled people if we restrict it to official social policy systems.

Both Australia and The Netherlands are characterized by more restricted and individualised eligibility for social benefits than in a less strict and more generous eligibility for social benefits in Norway – i.e., differences that potentially may trigger discontent or satisfaction in how people perceive themselves as fully included and recognised or excluded and mis-recognised in society [18]. Both Norway and Australia are heavily work-centered in their welfare and social policy (aiming for full employment) whereas The Netherlands focuses on a policy of social inclusion based on a more generic public rhetoric of participation in society with an emphasis on people’s well-being and community building [19].

To what extent and how potential differences in the organisation of public welfare provisions and the official rhetoric of inclusion correspond to differences in subjective experiences of inclusion for disabled people in daily life, however, is a more open question. National representative panel data from Australia in 2018 (N= 2,061, response rate = 71%) lend support to commonly held views of disabled people as easier to take advantage of, exploit, or treat badly, but also of an uncertainty as to how to act in an appropriate way towards disabled people. Even if this study reveals explicit or implicit negative stereotypical attitudes, and disempowerment, such as low expectations towards disabled people, it is equally interesting to note that 78% were unsure how to act towards disabled people, and that 42% agreed that people tend to ignore disabled people [12].

In line with Low and Pistaferri [18] we expect to find national differences in perceptions of inclusion/recognition between Norway on the one hand, and Australia and The Netherlands on the other hand, due to differences in disability and welfare systems and social policy. It may also be the case that more systemic features of society are only loosely associated with how people with an SCI perceive their inclusion and recognition in their own lives and as citizens. Sources of inclusion, and social recognition of a more informal kind (family, friends, and community) may play a significant role here.

Strictly speaking, our data do not fully cover the concept of social recognition, but focus on whether the respondents feel that they are treated with respect. Even if respect and recognition may differ theoretically in a larger philosophical discussion [9], there is also a substantial empirical overlap between being valued as a significant person and being respected as a member of society [20]. This provides the rationale for treating respect received from others as a measure of social recognition in this article.

### Research questions

RQ 1: To what extent are mobility limitations related to experiences of social inclusion/social recognition?

RQ 2: To what extent are mobility limitations an important determinant of whether people with an SCI feel included/socially recognised mediated by negative societal attitudes towards disabled people?

RQ 3: To what extent are differences in experiences of social inclusion/social recognition explained by country of residence?

The research questions are addressed with a focus on both factors explaining perceptions of inclusion and recognition, and factors explained by differences between societies.

### Hypotheses

Main Hypothesis: (1) Mobility limitations are related to experienced social inclusion and social recognition; (2) the association between mobility limitations and experienced social inclusion/social recognition is mediated by attitudes towards disabled people in society; (3) Levels of social inclusion and recognition factors associated with them vary across societies. These mechanism are presented schematically in Fig. 1:

**Fig 1.**
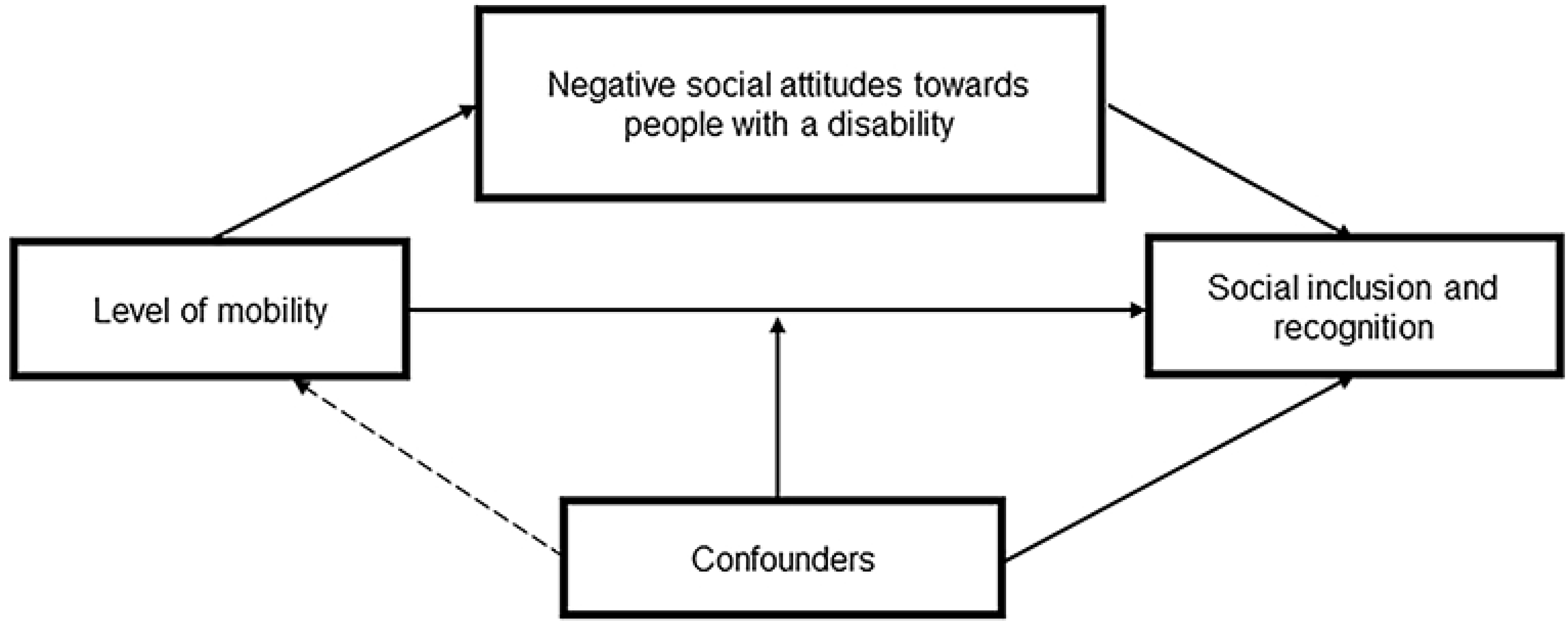
Schematic presentation of theoretichal mechanisms.

The model presents the theoretichal mechanisms in which level of mobility might affect social inclusion and recognition. The level of mobility of the respondent with SCI might have a direct effect on perceived levels of social inclusion and recognition. This relationship might be moderated by different confounders. The relationship might also be mediated by the experienced level of attitudes towards people with disabilities.

## Material and methods

### Data sources and sample

This study utilised data from the International Spinal Cord Injury (InSCI) Community Survey. The data derived from the core questionnaire with an extended national module on the attitudes and values of respondents from Norway, The Netherlands, and Australia [21–22]. The InSCI survey was conducted in Norway 19/10/2017 to 31/08/2018, in the Netherlands 01/01/2018 to 31/10/2018 and in Australia 01/03/2018 to 31/01/2019. The study embraced persons with SCI (traumatic or nontraumatic), aged 18 years or older, living in a community, and able to respond to one of the available language alternatives in which the 125-item questionnaire was made available. In Norway, potential participants were identified from the medical files of the three specialised SCI rehabilitation centres in the country (Oslo, Trondheim, and Bergen) who had completed their first rehabilitation in 2000-2016. This included 1,446 eligible persons and 611 respondents (response rate 42%). In The Netherlands, potential participants were identified from the medical files of three specialised rehabilitation centres in different parts of the country (Rotterdam, Utrecht, and Groningen). Random samples were drawn from the pools of all former patients in each centre. A total of 847 individuals were invited, and 260 (33%) participated in the study. In Australia, the data were based on specialised SCI clinical services/units, not-for-profit community organisations, and one government insurance agency in the states of New South Wales, Queensland, South Australia, and Victoria with specialised spinal cord services. A total of 5,925 were invited, and 1,579 participated in the study (i.e., a response rate of 27%).

### Ethics statement

The data collected in Norway, the Netherlands and Australia are all in compliance with national laws and regulatory approvals by Institutional Review Boards or Ethics Committees. All methods were performed in accordance with the Declaration of Helsinki. In Norway and the Netherlands, the participants provided their written informed consent by completing the survey (online or paper/pencil). In Australia implied consent was obtained from all participants by completing the survey questionnaire.

The Ethical Committees who approved the study were: Regionale komiteer for medisinsk og helsefaglig forskningsetikk (Regional committees for medical and health research ethics) in Norway (Reference number 2016/1184), Medish Ethische Toetsingscommisie (Medical Research Ethical Committee) in Utrecht, the Netherlands (Reference number 17/539) and Northern Sydney Local Health District, Australia (RESP/16/344).

### Sample description

The analytic sample consisted of 2,450 respondents. The Australian sample [23] consisted of over twice as many respondents as the Norwegian and the Dutch sample combined. Thus, the values of the total sample were significantly affected by the Australian sample. We took this into account, however, by either running the models separately for each of the three countries or applying the countries as dummy variables.

### Dependent variables

Both social inclusion and social recognition are elusive concepts difficult to measure and operationalise. To best capture the different nuances, we utilized two variables that were thought to measure inclusion and recognition, respectivley. The first dependent variable *Inclusion* asks the respondents “Do you feel included when you are with other people?” on a scale from 1 as *not at all*, up to 5 as *completely.* The proportion who do not feel included (1-2) was higher for The Netherlands (12%) and Australia (13%) respondents than in Norway (5%). The Netherlands and Australia followed each other to some extent in the distribution, whilst the Norwegians had more respondents placed higher up on the scale. For Norway, almost half of the sample (48.7%) felt completely included in the company of others, compared to one-third in The Netherlands (33.3%) and Australia (31.8%).

In order to measure the extent to which the respondents feel recognised, we apply the Recognition scale, which ask “do you feel that people treat you with respect?”. Here, the respondents place themselves on a scale from 1 to 7 where 1 is *not at all* and 7 is *a great deal*.

We observed a similar pattern for recognition as we did with how people felt included. 41.5% of Norwegian respondents reported that they got a great deal of respect (value 7), compared to 13.3% in The Netherlands and 28.1% in Australia. As seen in Table 1 and Table S1, most of the respondents in all three societies felt that they were included and recognised by others, but with a significantly higher proportion in Norway (8:10) than in The Netherlands and Australia (6:10).

**Table 1.** Descriptive statistics displaying the dependent variables and the main explanatory variables. a.

|  | Norway<br>(n= 517) | The Netherlands<br>(n= 236) | Australia<br>(n= 1,374) | Total<br>(N= 2,127 ) |
| --- | --- | --- | --- | --- |
| **Dependent variables** |  |  |  |  |
| Included (range 1-5) (mean, sd) | 4.25 (0.90) | 3.85 (1.08) | 3.79 (1.11) | 3.91 (1.08) |
| Treated with respect (range 1-7) (mean, sd) | 6.09 (1.00) | 5.52 (1.13) | 5.59 (1.32) | 5.70 (1.25) |
| **Independent variables** |  |  |  |  |
| Independence in Mobility |  |  |  |  |
| Electrical wheelchair (%) | 10% | 17% | 26% | 21% |
| Manual wheelchair (%) | 17% | 31% | 35% | 30% |
| Walking with devices (%) | 28% | 26% | 20% | 23% |
| Walking without devices (%) | 45% | 26% | 20% | 26% |
| Attitudes barriers |  |  |  |  |
| No influence/not applicable (%) | 82% | 73% | 72% | 74% |
| Made life harder (%) | 18% | 27% | 28% | 26% |

a. The number of respondents for each country (Norway, The Netherlands, and Australia) and the total number of respondents. The mean and standard deviation are presented for both of the dependent variables and percentages for each of the levels of mobility and perceived social attitudes.

### Explanatory variables

#### Independence in mobility

Mobility limitations were divided into four different categories. The first category included respondents who were completely dependent on help from others or use an electric wheelchair. Respondents in the second category were those using a manual wheelchair. Those in the third and fourth category could walk moderate distances with help from walking aids. Half of the respondents in this study were dependent on a wheelchair (21% electric wheelchair users vs. 30% manual wheelchair users), one out of four (23%) could walk short distances with walking aids, and one out of four (26%) were able to walk short distances without walking aids. The proportion of participants depending on wheelchair use was substantially higher in Australia (60%) than in The Netherlands (38%) and Norway (27.5), and conversely lower for people able to walk (with or without walking aids) in Australia (40%) than in The Netherlands (62%) and Norway (72.5%).

#### Negative attitudes towards people with a disability in society

People with SCI are exposed to various external influences or environmental factors in daily life. A question on social attitudes was included in the survey regarding these environmental factors, namely “Thinking about the <u>last 4 weeks</u>, please rate how much these environmental factors have influenced your participation in society.” Participants rated this item on a scale of 4, where 1 = Not applicable, 2 = No influence, 3 = Made my life a little harder, and 4 = Made my life a lot harder.

These four categories were recoded as a dichotomous variable coded as “0” if not applicable or no influence, and “1” if societal attitudes have made life a little harder or a lot harder. Two to three out of ten feel that negative attitudes towards disabled people have made life more difficult for them (18% in Norway, 26% in The Netherlands, and 28% in Australia).

#### Control variables

As control variables, we included gender, age group (based on age at the time of interview), level of education, and job status. Female was coded as 1, male was coded as 0. Age was a categorical variable. The level of education was coded as ”0” with elementary schooling; “1” with completed secondary education; “2” with completed university, whether they went on to graduate study or not. The final control variable measured whether the respondents had been in remunerative work during the previous seven days (no= 0, yes= 1). Thus, respondents who were still enrolled in school, retired, or for some reason unemployed were categorised/classified as not in paid work.

We run two regressions utilising *ordinary least square regression*. The first explored the relative impact of mobility limitations on experiences of social inclusion/social recognition (research question 1) and its impact after controlling for negative societal attitudes towards disabled persons (research question 2). The second focused on experiences of social inclusion and recognition explained by country of residence.

## Results

The results presented in Figure 2 lend limited and uneven support for mobility limitation as a factor explaining experiences of social inclusion/social recognition (cf. research question 1).

**Fig 2.**
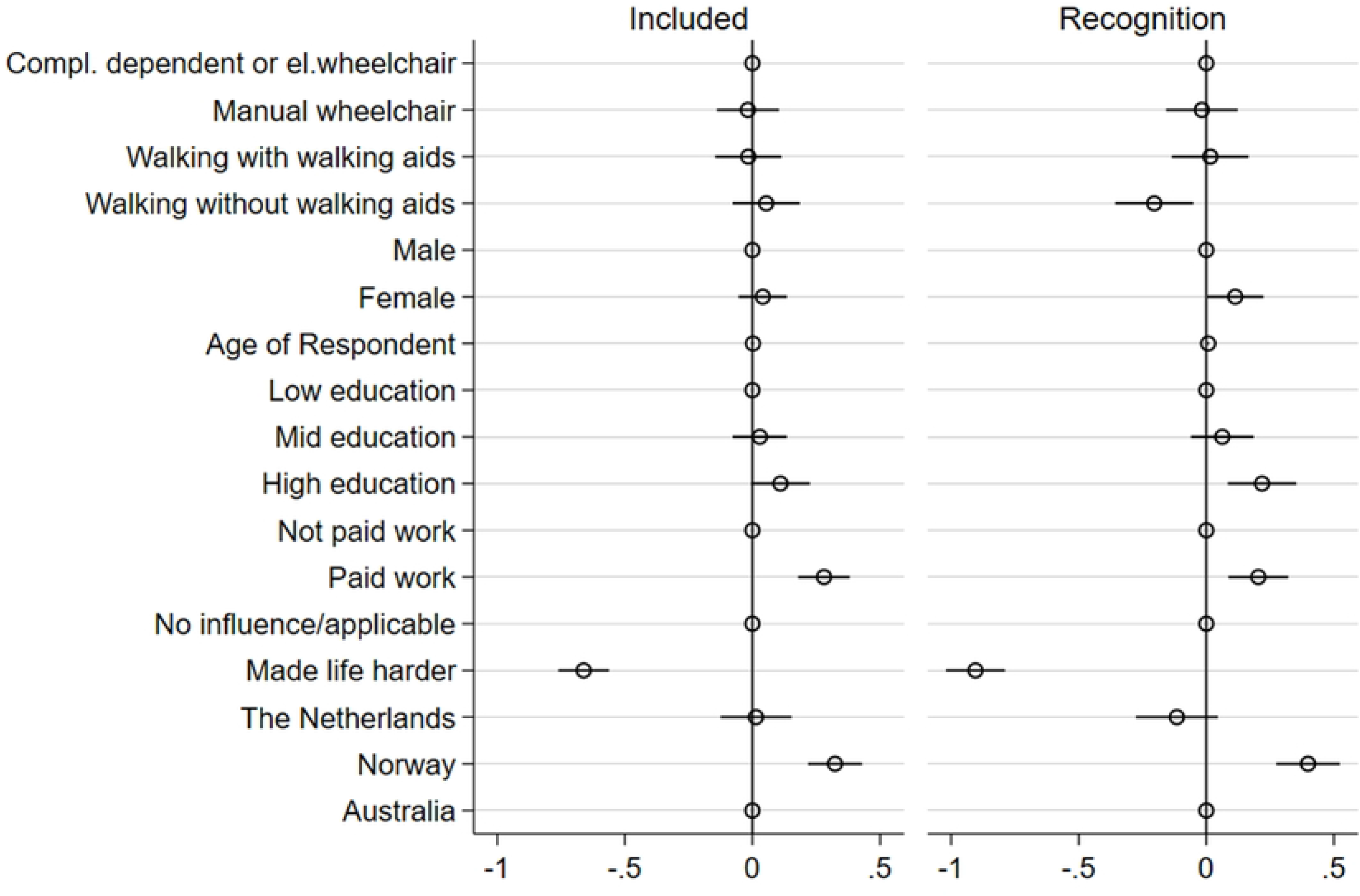
Plot comparing regression coefficients on the dependent variables included and Recogniced controlling for countries.

The plot is based on Table S2. It shows regression coefficients on the effect on the experience of being included and the experience of recognition. The figure included the reference category for each dummy set (reference categories are: completely dependent of wheelchair or using an electric wheelchair, male, low education, not paid work, no influence/applicable, and The Netherlands).

The relative impact of negative societal attitudes towards disabled people is overall a much stronger predictor of how people with an SCI experience social inclusion/social recognition (cf. research question 2).

Walking without walking aids and age may have a positive effect on feelings of being included in company with others but disappear as an effect as soon as we control for negative attitudes towards people with a disability. Recognition, however, appears to be a more complex case where those walking without walking aids report less respect from others than those dependent on a wheelchair. Figure 2 also suggests that women with an SCI more often feel accorded respect than men, and those who are older more often feel accorded respect than those of a younger age.

The results in figure 2 show that recognition is strongly linked to higher education and being employed. It also reveals that Norwegian respondents report overall better results on inclusion and recognition than respondents in Australia and The Netherlands.

With regard to the mediation effects of social attitudes, the results are mixed. Environmental factors of social attitudes have a negative and significant effect on both inclusion and recognition. Inclusion of social attitudes affects the p-value of the main explanatory variablie of mobility, going from significant to not significant. This suggests that there might be som mediating effect of social attitudes on the relationship between mobility and inclusion. In terms of recognition, walking without aids becomes significant and negative in relation to the being completely dependent on an electric wheelchair. However, since the relationship between mobility and recognition did not have a significant association prior to the inclusion of social attitudes, we cannot show a potential mediation effect in our study. Figure 3 and Figure 4 presents the coefficient plots for both inclusion and recognition for each of the three countries.

**Fig 3.**
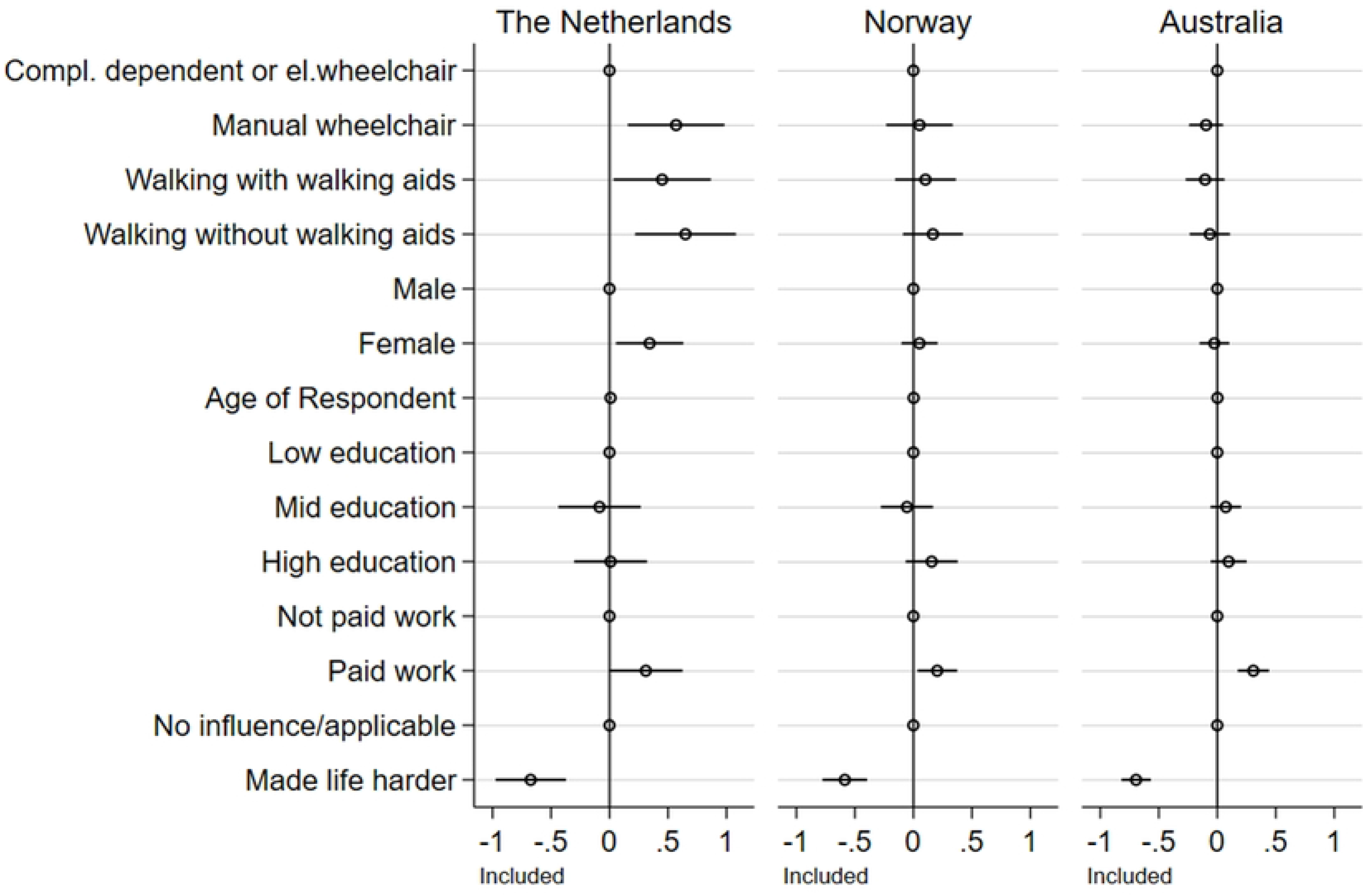
Regression coefficients. Dependent variable: Inclusion. The plot is based on Table S3. It shows regression coefficients on the effect on the experience of being included for The Netherlands, Norway, and Australia. The figure includes the reference category for each dummy set (reference categories are: completely dependent of wheelchair or using an electric wheelchair, male, low education, not paid work, no influence/applicable, and The Netherlands).

**Fig 4.**
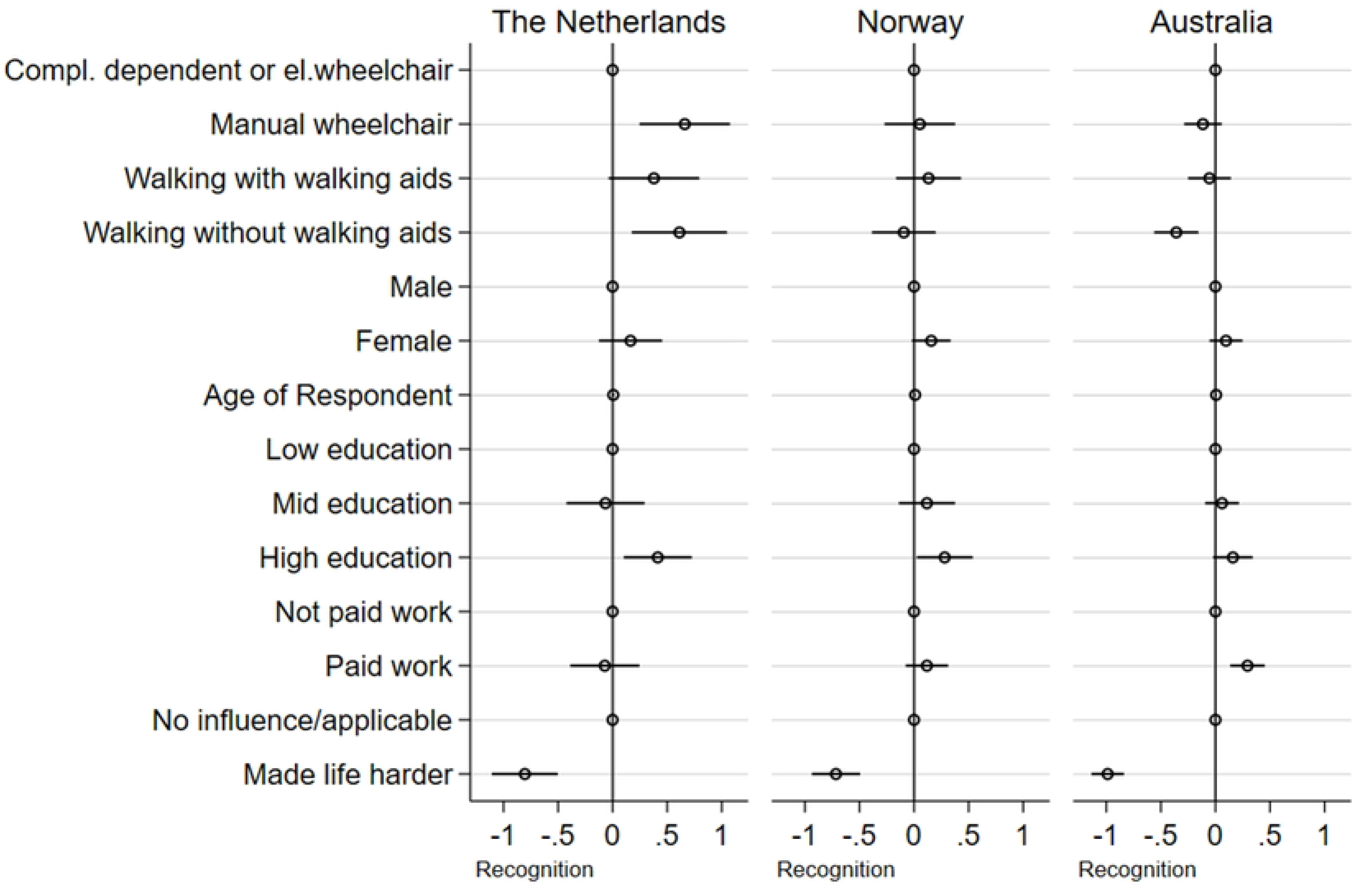
Regression coefficients. Dependent variable: Recognition. The plot is based on Table S3. It shows regression coefficients on the effect on the experience of recognition for The Netherlands, Norway, and Australia. The figure includes the reference category for each dummy set (reference categories are: completely dependent of wheelchair or using an electric wheelchair, male, low education, not paid work, no influence/applicable, and The Netherlands).

Our initial hypothesis that mobility limitations may impact experiences of social inclusion/social recognition received some support in The Netherlands where both manual wheelchair users and those walking without walking aids reported higher levels of social inclusion and social recognition. An impact on social recognition of walking without walking aids was also found in Australia, albeit as a negative factor. This hypothesis received no support in the Norwegian data.

For both Norway and Australia, it was observed that the participants’ sense of inclusion is heavily dependent on their status as employed or not. In The Netherlands it is primarily gender, in addition to mobility status, not employment, that explains differences in how people with an SCI perceive their inclusion in society.

In Norway, it was observed that gender, age, and education play a role in how the respondents report being recognised by others, whereas gender disappears after introducing negative attitudes towards disabled people in society. In The Netherlands, recognition is primarily associated with mobility status, age, and education, after controlling for negative attitudes towards disabled people. In both Norway and The Netherlands having a high education compared to lower education is positively associated with recognition. This is, however, not the case in the Australian sample. In addition to mobility status, and paid work, older people in Australia were more often report that they felt socially recognised than people of a younger age. Interestingly, walking without aids in the Australian sample, negatively associated with recognition compared to being completely dependent, having a different effect compared to The Netherlands.

In summary, these results show that both explanations departing from impairment and mobility constraints (i.e., constraints typically represented in the rehabilitation literature) and discrimination towards disabled people in society are robust factors in explaining social inclusion and social recognition. At the same time, we also find a more complex picture than initially expected.

## Discussion

We initially started out with a hypothesis (H1) that mobility limitations are related to self-perceived experienced social inclusion and social recognition (see also research question 1). This was based on an interest in how physical mobility constraints impact social participation with a focus on both perceptions on individual inclusion and social recognition from other people. It was also based on an interest in whether it is physical mobility, or the symbolic signs associated with a disability that matter most in how people with a physical impairment perceive themselves. We find some support in both the Dutch and the Australian data that walking without walking aids may have an impact on social recognition, albeit in opposite directions from those depending on an electric wheelchair.

Whereas no visible signs of a prototypical impairment may be an asset, signaling “normality” and “sameness”- i.e., no deviance in public, it may also represent a challenge. This is particularly so if we depart from a social recognition perspective, where an initial impairment is less visible and obvious, and therefore potentially mis-recognised as a constraint and personal challenge in the public sphere. This is also personal challenges that has been described in previous studies in other minor groups, including minor stroke [24], chronic fatigue [25] and pain [26].

It is also interesting to observe that the cultural marker in The Netherlands is not necessary between wheelchair users and those able to walk without walking aids but associated with mobility status (where both those able to walk and those sitting in a manual wheelchair more often report that they are socially recognised than in those in an electric wheelchair). The weak correlations between mobility status and social recognition are also an illustration of a factor highly dependent on society, culture, and attempts to strive for a culture of “normality” and a discourse of disability [27].

We expected to find an empirical correlation between mobility limitations and individual perceptions of inclusion/social recognition mediated by negative attitudes towards people with a disability (H2). Even if this hypothesis receives some support in The Netherlands and Australia, the overall strongest predictor of both social inclusion and social recognition is negative attitudes towards disabled people. Our answer to research question 2 is accordingly that negative encounters of a personal or more generic kind towards disabled people are essential in any study of social inclusion and social recognition. This is also a result in line with a broad range of disability research [3, 6] and more general literature on social recognition as a key to understand social inclusion [11, 9, 28]. The main difference of the approach taken in this article from what may be found in the literature above is that we focus upon experiences of inclusion and recognition based on extensive quantitative data from people with an SCI rather than more general domains of participation and inclusion in society.

We expected to find differences in how people with an SCI experienced social inclusion and social recognition across the three societies (cf. H3, and research question 3). This was based mainly on more general assumptions of societal traits found in social welfare and social policy literature where both Australia and The Netherlands typically (and in line with Canada, the United Kingdom, and the USA) are described as more liberal, consumer-, and market-oriented in their approach towards disability and health. In contrast to both Australia and The Netherlands we initially expected to find overall better results in terms of inclusion, recognition, and negative reporting of discrimination of people with a disability in Norway. This was also an idea based on our knowledge of differences in means testing of disability aid and welfare services, potentially associated with more stigma in both Australia and The Netherlands than in more comprehensive unconditional means testing programs in Norway. The data lend some support for a fit between type of society and how people with a disability perceive their inclusion and recognition as persons in a society, where Norway ends up with overall better results than either Australia or The Netherlands. It is nonetheless difficult to interpret individual behavior as a response to differences in welfare social policy and disability policy without considering alternative sources of inclusion and recognition that may compensate for institutional and more systemic limitations and constraints.

Norway may on paper be more disability-friendly based on its commitment to personalised services but does not necessarily score as well on social capital in the community and assistance from family and friends. Australia scores overall surprisingly low on disability-friendly measures in an OECD context [17] but may also have the relative advantage of a more community- and family-based model of inclusion. The Netherlands has overall higher scores on disability-friendly measures in an OECD context than in Australia [18] but its differences in terms of mobility status and the significance of prototypical disability markers mark it out as taking a distinct path from Norway and Australia. To what extent this may be explained by a more pronounced disability discourse based on individual identity management and performativity [2, 3]) or moral imperatives associated with inclusion and expectations to strengthen the well-being of those with disabilities [19] remains to be researched. This is also a question of more general interest as the results for The Netherlands in contrast to both Norway and Australia, show a weak link to social inclusion and social recognition via employment as the main road to both inclusion and recognition in society.

### Study Limitations

The study was limited in size, and with an uneven response rate. Ideally, we would have liked to have included more countries and countries representing a broader range of health and welfare systems. All data are based on self-reporting, not allowing for full validation of the data used. As noted, the differences in the final sample within each country produced some limitations concerning the comparability between the three countries. Whilst Norway and The Netherlands, to some extent, consisted of a similar sample, the Australian sample differed slightly. For instance, a higher share of the respondents in the Australian sample had a complete level of SCI compared to the Norwegian sample. In addition, the average time since the onset of SCI was twice as high in Australian, again, compared to the Norwegian sample. Taken together, these variations could lead to biased results.

The measures used are proxies for the concepts we wanted to measure. The question about being treated with respect by others may be regarded as a more individualised construct than social recognition, which is a broader sociological category also including the social bonds and the solidarity that constitutes groups, communities, and society. Our use of *respect,* however, is primarily framed in sociological language as our focus is on social and societal factors impacting how people with a disability perceive themselves in different societies.

## Conclusions

Our departure in this article differs from the approaches taken in previous literature in several respects. First, the article highlights and problematises the nexus between mobility limitations in everyday life and perceptions of inclusion and social recognition. Second, we did not restrict our research to one society but examined how this nexus is played out in three distinct societies, with data from Norway, The Netherlands, and Australia.

Our results lend some support to a general assumption that a focus on mobility limitations may have an impact on how people with an SCI perceive themselves, including whether they consider themselves included and fully recognised by others in society. This is, however, a rather limited focus if we compare it to the challenges people with an SCI face when it comes to negative attitudes and stereotypical attitudes in Norway, The Netherlands, and Australia. In line with this, it is obvious that both people with an SCI and their relative’s post-SCI must cope with and adjust to the challenges associated with disability in society.

The article confirms that there are significant physical barriers and obstacles for people with an SCI in all three nations to move around due to the degree of impairment, i.e., problems traditionally addressed in disability and equal access policy, and in a vocabulary of accessible, usable and universal design (see also United Nations – Article 9 on Accessibility as a mechanism to secure social inclusion in the UN Convention of Human rights for people with a disability [29]).

At the same time, we find an even stronger social and cultural dimension linked to societal factors in attitudes linked to social recognition and perception of Self and Others. This may not come as a big surprise but is interesting as it must be read considering different welfare regimes and national policy contexts, and ways to handle disability in social policy and practice. It is also of interest, as we move from a broader societal context to an interest in actors and social interactions between people. Once we bring in the social interaction and social space, we also observe that mobility limitations are not just a matter of physical constraints, but also a question of how a wheelchair or mobility aids, associated with different sets of cultural stereotypes, can function. Rather than being an asset, our results suggest that an SCI without the necessary stereotypical attributes (read dependent on a wheelchair) may be an obstacle in terms of being fully recognised as a person.

In a context of recognition, and social inclusion in society, people with a disability are not just of interest as individuals or a group, but also in terms of their social roles and resources. This is particularly important in an inquiry about social recognition. Societal roles linked to work and employment, and resources such as educational capital, are to a high extent status markers and social distinctions of importance how individuals see and perceive themselves, and how they are seen and perceived by others [9, 20].

The results support previous research that both higher education and employment are essential in how people with an SCI perceive themselves and are perceived by others [27]. The results also lend support to a picture where women with an SCI more often feel accorded respect than men, and those older more often feel accorded respect than those of a younger age. This fits rather well with a theoretical understanding of social recognition as norm structures and systems that are more heavily embedded in the cultural discourse for men compared to women, and amongst younger compared to elderly. People age 50+ have also in previous SCI research in Norway been found to be more prone to say that they feel that they have done their societal duty as citizens based on their previous work careers and track records [27].

The results lend some support to identifying commonalities across societies, but also to specifying national differences with respect to the importance of employment and higher education as social status and social recognition markers/distinctions. The results from The Netherlands are of particular interest as they appear to support a narrative of inclusion and recognition via a general idea of social participation and capability, in contrast to more conventional status markers as higher education and employment in Norway and Australia.

More research is needed to fully understand the interplay between self-perceived experiences of being included and recognised by others, and a more generalised recognition of people with a disability in society. Our results are, nonetheless, a strong reminder that health professionals in charge of the rehabilitation of people with an SCI must pay more attention to societal factors and social barriers associated with disability. We also found confirmation of the notion that it is not necessarily mobility limitations that are the main challenge in everyday life, but rather the stereotypical attitudes and norms people encounter. Stereotypical attitudes and norms in society are also shown to have diverse impacts on recognition in the three countries.

Attitudes (personal as well as more generalised attitudes) are not in themselves a mirror of how people behave or act. Attitudes interpreted as prevailing value systems in societies, may also be of a more ambivalent and contradictory nature (in support of official hegemonic norms, yet ambivalent or not fully considered as in the case of public attitudes towards disabled people [12]. Nonetheless, we argue that an international comparative survey addressed to disabled people is crucial for our understanding of commonalities and variations in how social recognition is lived and perceived for people with a spinal cord injury.

## Data Availability

Data availability: See InSCI. 2017. “International Spinal Cord Injury Community Survey – What is InSCI?”. URL: https://insci.network/insci/T1/en/what_is_insci.php. In SCI data are available on request. For more information about the dataset used to analyse the material presented in the paper, please contact Annelie S. Leiulfsrud at Norwegian University of Science and Technology (NTNU) by.

## Supporting information

**S1 Table. Results on the questions on “social inclusion” and “people respect” in Norway, The Netherlands and Australia.**

**AS2 Table. Regression effect on perceived inclusion and recognition.** Controlling for country.

**S3 Table. Regression table for the effect on perceived inclusion and recognition for Norway, The Netherlands, and Australia.**

## References

1. Ridolfo H, Ward BW. Mobility impairment at the construction of identity. First Forum Press. Boulder, Co; 2013.

2 . Grue J. The social meaning of disability: a reflection on categorisation, stigma and identity. Social Health and Illness. 2016; 38(6): 957–964. 10.1111/1467-9566.12417

3. Darling RB. Disability and identity – Negotiating self in a changing society. Lynn Rienner Publishers. Boulder. Co; 2019.

4 . Cahill SE, Eggleston R. Reconsidering the stigma of physical disability, Wheelchair Use and Public Kindness. The Sociological Quarterly. 2016; 36(4): 681–698. 10.1111/j.1533-8525.1995.tb00460.x

5 . Osterthun R, Postma K, van den Berg RHJ, Stolwijk J, Tepper M, Post MWM. Hoe ervaren mensen met verschillende mobiliteit na een dwarslaesie hun gezondheid, functioneren en welbevinden? [Associations between mobility level and levels of health, functioning and well-being]. Ned Tijdschr Revalidatiegeneeskd 2021; 43(2): 32–36. https://www.revalidatie.nl/wp-content/uploads/2023/04/NTR-2021-2.pdf

6 . Oliver M, Barnes C. Back to the future: the world report on disability. Disability and Society. 2012; 27(4): 575–579. 10.1080/09687599.2012.686781

7 . Schirghuber J, Schrems B. Being wheelchair-bound and being bedridden: Two concept analyses. Nursing Open. 2023;10(4): 2075–2087. 10.1002/nop2.1455

8 . Sapey B, Stewart J, Donaldsen G. Increases in wheelchair use and perceptions of disablement. Disability and Society. 2005; 20(5): 489–505. 10.1080/09687590500156162

9 . Honneth A. Recognition and justice: Outline of a plural theory of justice”. Acta Sociologia. 2004; 47(4): 351–364. 10.1177/0001699304048668

10. Ricouer P. The Course of Recognition. Cambride. Havard University Press; 2007.

11 . Fraser N. Rethinking recognition. New Left Review; 2003; 3: 107–120.

12 . Bollier AM, Sutherland G, Krnjacki L, Kasidis V, Katsikis G, Ozge J, et. al. Attitudes Matter: Findings from a national survey of community attitudes toward people with disability in Australia. Centre of Research Excellence in Disability and Health. The University of Melbourne. 2021. doi: 10.26188/15176013

13. OECD. Trust in government (indicator). 2022 doi: 10.1787/1de9675e-en. Available from: https://www.oecd-ilibrary.org/sites/b407f99c-en/index.html?itemId=/content/publication/b407f99c-en

14. Esping-Andersen G. The three worlds of welfare capitalism. Princeton, New York: Princeton University Press; 1990.

15 . Headey B, Goodin RE, Muffels R, Dirven HJ. Welfare Over Time: Three Worlds of Welfare Capitalism in Panel Perspective. Journal of Public Policy. 1997; 17(3): 329–359. 10.1017/S0143814X00008576

16. Aspalter C. Different Worlds of Welfare Capitalism: Australia, the United States, the United Kingdom, Sweden, Germany, Italy, Hong Kong, and Singapore. Australian National University Discussion Paper. 2010, December. (80). Available at SSRN: https://ssrn.com/abstract=1725128

17. OECD. Sickness, disability, and work. Breaking the barriers. Norway, Switzerland and Polen. Paris. OECD. 2010. Available from: https://read.oecd-ilibrary.org/social-issues-migration-health/sickness-disability-and-work-breaking-the-barriers_9789264088856-en#page1

18 . Low H, Pistaferri L. Disability Insurance and the Dynamics of the Incentive Insurance Trade-Off. American Economic Review. 2015; 105(10): 2986–3029. DOI: 10.1257/aer.20110108

19. Freitas MJ, Dassen J, Louali S, Sniekers M, Van Lieshout C, Wevers C. Inclusive Society and Social Work: The Netherlands. In: Bundschuh S, M.J Freitas MJ, Bartroli CP, Žganec N. (eds) Ambivalences of Inclusion in Society and Social Work. European Social Work Education and Practice. Springer, Cham; 2021

20. Sayer A. Class, Moral Worth and Recognition. London, Thousand Oaks, New Delhi. SAGE Publications; 2005.

21 . Gross-Hemmi MH, Post MWM, Ehrmann C, Fekete C, Hasnan N, Middleton JW, et. al. Study protocol of the international Spinal Cord Injury (InSCI) Community Survey. American Journal of Physical Medicine and Rehabilitation. 2017; 96(2): 523–534. 10.1097/phm.0000000000000647

22 . Fekete C, Brach M, Ehrmann C, Post MWM, InSCI, Stucki G, et. al. Cohort Profile of the International Spinal Cord Injury Community Survey Implemented in 22 Countries. Archives of Physical Medicine and Rehabilitation. 2020; 101:12103–2111. doi: 10.1016/j.apmr.2020.01.022

23 . Middleton JW, Arora M, Kifley A, Geraghty T, Borg SJ, Marshall R, et.al. Australian arm of the International Spinal Cord Injury (Aus-InSCI) community survey: 1. population-based design, methodology and cohort profile. Spinal Cord. 2023 Mar;61(3):194–203. 10.1038/s41393-022-00850-6

24 . Zheng Z, Song R, Zhao Y, Lv H, Wang Y, Yu C. 2023. An investigation of the stigma and the factors influencing it in the rehabilitation of young and middle-aged stroke patients – a cross-sectional study. BMC Neurology 23: 139. 10.1186/s12883-023-03189-4

25 . Froehlich L, Hattesohl DB, Cotler J, Jason LA, Scheibenbogen C, Behrends U. Causal attributions and perceived stigma for myalgic encephalomyelitis/ chronic fatigue syndrome. Journal of Health Psychology. 2022; 27(10): 2291–2304. 10.1177/13591053211027631

26 . De Ruddere L, Craig KD. Understanding stigma and chronica pain: a-state-of the-art review. Pain. 2016; 157(8): 1607–1610. 10.1097/j.pain.0000000000000512

27. Leiulfsrud AS. Exploring Persons With a Spinal Cord Injury Participation in Society: The Paradoxes of the Participation Dimension in the International Classification of Functioning, Disability and Health (ICF). Doctoral Thesis NTNU 2016:324.Norwegian University of Science and Technology, Trondheim, Norway. ISBN: 978-82-326-1987-0

28. Gimmler A. Recognition: Conceptualization and context. In: Leiulfsrud H, Sohlberg P (Eds). Concepts in Action. Leiden, Boston. Brill; 2018.

29. United Nations – Article 9. Accessibility. Available from: https://social.desa.un.org/issues/disability/crpd/article-9-accessibility

